# Citation screening using large language models for creating clinical practice guidelines: A protocol for a prospective study

**DOI:** 10.1101/2023.12.29.23300652

**Authors:** Takehiko Oami, Yohei Okada, Taka-aki Nakada

## Abstract

**Background:** The development of clinical practice guidelines requires a meticulous literature search and screening process. This study aims to explore the potential of large language models in the development of the Japanese Clinical Practice Guidelines for Management of Sepsis and Septic Shock (J-SSCG), focusing on enhancing literature search quality and reducing the citation screening workload.

**Methods:** A prospective study will be conducted to compare the efficiency and accuracy of literature citation screening between the conventional method and a novel approach using large language models. We will use the large language model, namely GPT-4, to conduct literature searches for predefined clinical questions. We will objectively measure the time required for citation screening and compare it to the time taken using the conventional method. Following the screening, we will calculate and compare the sensitivity and specificity of the results obtained from the conventional method and the large language models-assisted process. The total time spent using both approaches will also be compared to assess workload reduction.

**Trial registration:** This research is submitted with the University hospital medical information network clinical trial registry (UMIN-CTR) [UMIN000053091].

**Conflicts of interest:** All authors declare no conflicts of interest to have.

**Funding:** None

## Background

The development of clinical practice guidelines is crucially dependent on systematic reviews to gather and compile the latest research evidence. This fundamental but burdensome process involves repetitive citation screening, a task that is notoriously time-consuming and labor-intensive [1-3]. Recent advancements have seen reports on machine learning applications for citation screening, offering hopes of workload reduction [4-9]. However, previous findings from our own research have only shown time savings without achieving satisfactory accuracy. This underscores the necessity for more efficient methods that further refine the accuracy of citation screening [9-11].

Amidst growing interest in large language models (LLMs), including ChatGPT, these advanced artificial intelligence tools, rooted in natural language processing, have shown the potential in performing complicated tasks such as data interpretation and text generation [12, 13]. This newly launched technology is a promising candidate to transform the whole process of citation screening owing to its capacity of generating human-like responses based on highly sophisticated comprehension [14, 15]. As an example of this offer, previous research has suggested the feasibility of utilizing GPT-3.5 for citation screening tasks [16]. Yet, there remains a dearth of studies focused on the application of LLMs for extensive citation screening in the context of developing clinical practice guidelines.

Therefore, we hypothesize that a large language model may potentially meet the quality of manual citation screening and substantially diminish the manual workload. This prospective study will critically compare the accuracy and operational efficiency of LLMs with that of conventional manual screening methods, with the aim of setting a new standard in the creation of clinical practice guidelines.

## Methods

### Study design and settings

We will conduct a prospective study to validate the efficacy of LLMs for citation screening. This research will involve several phases: the elaboration of a suitable query for LLMs based on the prompt engineering technique [17][18], setting of execution commands to complete the subsequent tasks of LLMs, and an automated implementation of the citation screening process. To ensure transparency and reproducibility, we have documented our review protocol and disseminated it through the medRxiv pre-print server. Additionally, our study is registered with the University Hospital Medical Information Network (UMIN) clinical trials registry under the identifier [UMIN000053091], underscoring our commitment to rigorous scientific standards and ethical research practices.

### Clinical questions in the J-SSCG

We will evaluate the efficiency of the large language model using the clinical questions (CQs) in the revised version of J-SSCG 2020, namely J-SSCG 2024, as described in the previous report. The Japanese Society of Intensive Care Medicine (JSICM) and the Japanese Association for Acute Medicine (JAAM) created J-SSCG 2020 and will publish J-SSCG2024, in light of Japanese-specific clinical settings for sepsis and septic shock on the clinical practice [19].

The five CQs (Table 1), which we deployed in the previous report, will be used for this study. Briefly, extensive literature reviews were carried out via CENTRAL, PubMed, and Ichushi-Web for these CQs, with the working group members ensuring a comprehensive search strategy that includes all key studies. The literature was limited to Japanese and English. All titles and abstracts were downloaded, collated, and deduplicated using EndNote as the citation manager for J-SSCG 2024.

**Table 1.** The list of the patient/population/problem, intervention, and comparison of the selected clinical questions.

|  | Patient, population, problem | Intervention | Comparison |
| --- | --- | --- | --- |
| CQ1 | Adult patients (18 years old or older) diagnosed with or suspected of having infection, bacteremia, or sepsis | Balanced crystalloid administration | 0.9% sodium chloride administration |
| CQ2 | Adult patients (18 years old or older) with sepsis, or suspected as sepsis, infection, bacteremia or patients admitted to ICU | Targeting a higher mean arterial pressure | Targeting a lower mean arterial pressure |
| CQ3 | Adult patients (18 years old or older) with sepsis presenting with severe metabolic acidosis or patients admitted to ICU | Sodium bicarbonate administration | No sodium bicarbonate administration |
| CQ4 | Adult patients (18 years old or older) with sepsis or septic shock | Usual care with at least one of the following tissue perfusion parameters: lactate/lactate clearance, capillary refill time, ScvO<sub>2</sub>/SvO<sub>2</sub>, and P(v-a) CO<sub>2</sub>/C (a-v) O<sub>2</sub>. | Usual care with different parameters mentioned in the interventional group, or standard care without the utilization of any specific tissue perfusion parameters |
| CQ5 | Adult patients (18 years old or older) with sepsis, sepsis-induced hypotension, or septic shock | Restrictive fluid management, which aims to reduce the amount of fluid therapy for up to 24 h | Conventional fluid management or non-restrictive fluid management defined by authors |

CQ: clinical question; ICU: intensive care unit

### Large language model

We will use GPT-4 as a large language model to investigate the efficiency and feasibility for implementing systematic reviews in the development of clinical practice guidelines. After importing the dataset from citation managers with the same procedure of the conventional tool for citation screening, we interacted the dataset with OpenAI GPT4 API using pandas (v1.0.5) in Python (v3.9.0). To conduct ChatGPT-guided systematic reviews, we created a query to have ChatGPT automatically implement a citation screening process as follows. As of each query, we will follow the exactly same phrases described in the framework of CQs that the J-SSCG2024 members of the conventional citation screening created.

Prompt:

You are conducting a systematic review and meta-analysis, focusing on a specific area of medical research. Your task is to evaluate research studies and determine whether they should be included in your review. To do this, each study must meet the following criteria:

Target Patients: ---

Intervention: ---

Comparison: ---

Study Design: ---

Additionally, any study protocol that meets these criteria should also be included.

However, you should exclude studies in the following cases:

The study does not meet all of the above eligibility criteria.

The study’s design is not a randomized controlled trial. Examples of unacceptable designs include case reports, observational studies, systematic reviews, review articles, animal experiments, letters to editors, and textbooks.

After reading the title and abstract of a study, decide whether to “include” or “exclude” it based on these criteria.

Title: ---

Abstract: ---

Then, we will implement the citation screening using the following code. According to the relevance of the references, GPT-4 will decide to include or exclude each citation without prior knowledge. After the session, we will check the results recorded on the file. The code for this process will be available at the following link: https://github.com.

### Conventional citation screening

We will use the same data for the conventional citation screening in the previous data published as a prospective observation study. Briefly, EndNote-processed files was transferred to Rayyan, a systematic review assistant software. Two independent reviewers screened the studies by title and abstract, resolving conflicts through discussion or third-party review.

### Data collection

Our research will compare the accuracy and workload between traditional literature screening methods and those assisted by a large language model. To assess accuracy, we will count the number of missing or redundant references identified by the conventional screening as compared to the large language model-assisted process. To assess workload, we will record the time taken to citation screening. The reviewers will use a command line to measure the processing time to complete the task. For the conventional screening, we will use the previously published data that recorded the duration of the screening session using Rayyan’s built-in time tracking feature.

### Statistical analysis

Continuous variables will be reported as means with standard deviations or medians with interquartile ranges, based on their distribution characteristics. The efficacy of the LLM in systematic reviews will be evaluated by comparing its sensitivity and specificity with the conventional method, which will serve as the benchmark for accurate citation screening. Literature identified as “relevant” by GPT-4 will be considered for inclusion. To calculate sensitivity, specificity, and positive predictive value in the primary analysis, we will use the second screening results in the conventional method as the gold standard to compare the results of the first screening performed by both GPT-4 and the conventional method. The results of the first screening session using the conventional method will serve as the gold standard in the secondary analysis to evaluate the performance of the screening process facilitated by GPT-4 in terms of the statistical measures. The duration of systematic review sessions for each clinical question will be collectively analyzed to measure time efficiency. As a sensitivity analysis, the study will analyze the variations in accuracy attributable to the modifications of the GPT-4 prompts, thereby examining how prompt engineering influences LLM performance in citation screening tasks. For statistical analysis, the GraphPad Prism 10 software (GraphPad Software, San Diego, CA, USA) will be used to manage the research data and perform statistical tests.

## Data Availability

All data produced in the present study are available upon reasonable request to the authors.

## Notes

### Competing Interest Statement

The authors have declared no competing interest.

### Clinical Protocols

https://www.umin.ac.jp/ctr/index-j.htm

### Funding Statement

This study did not receive any funding.

### Author Declarations

Since this research does not include the conditions as defined in "Life Science and Medical Research Involving Human Subjects", we consider this research to be outside the scope of research for which an application for ethical approval is required.

